# Diagnosing Cornelia de Lange syndrome and related neurodevelopmental disorders using RNA-sequencing

**DOI:** 10.1101/19008300

**Authors:** Stefan Rentas, Komal S. Rathi, Maninder Kaur, Pichai Raman, Ian D. Krantz, Mahdi Sarmady, Ahmad Abou Tayoun

**Affiliations:** Division of Genomic Diagnostics, The Children’s Hospital of Philadelphia, Philadelphia, PA, 19104, USA; Department of Biomedical and Health Informatics, The Children’s Hospital of Philadelphia, Philadelphia, PA, 19104, USA; Roberts Individualized Medical Genetics Center, Division of Human Genetics, The Children’s Hospital of Philadelphia, Philadelphia, PA, 19104, USA; Genomics Center, Al Jalila Children’s Specialty Hospital, Dubai, UAE

**Author notes:** These authors contributed equally to this work (Stefan Rentas, Komal S. Rathi). Correspondence, 1-267-426-1373. Correspondence, 971-05-193-3105.

**Keywords:** RNA-seq, lymphoblastoid cell line, Mendelian gene, neurodevelopmental disorder, Cornelia de Lange Syndrome, genetic testing

## Abstract

**Purpose:** Neurodevelopmental phenotypes represent major indications for children undergoing clinical exome sequencing. However, 50% of cases remain undiagnosed even upon exome reanalysis. Here we show RNA sequencing (RNA-seq) on human B lymphoblastoid cell lines (LCL) is highly suitable for neurodevelopmental Mendelian gene testing and demonstrate the utility of this approach in suspected cases of Cornelia de Lange syndrome (CdLS).

**Methods:** Genotype-Tissue Expression project transcriptome data for LCL, blood, and brain was assessed for neurodevelopmental Mendelian gene expression. Detection of abnormal splicing and pathogenic variants in these genes was performed with a novel RNA-seq diagnostic pipeline and using a validation CdLS-LCL cohort (n=10) and test cohort of patients who carry a clinical diagnosis of CdLS but negative genetic testing (n=5).

**Results:** LCLs share isoform diversity of brain tissue for a large subset of neurodevelopmental genes and express 1.8-fold more of these genes compared to blood (LCL, n=1706; whole blood, n=917). This enables testing of over 1000 genetic syndromes. The RNA-seq pipeline had 90% sensitivity for detecting pathogenic events and revealed novel diagnoses such as abnormal splice products in *NIPBL* and pathogenic coding variants in *BRD4* and *ANKRD11*.

**Conclusion:** The LCL transcriptome enables robust frontline and/or reflexive diagnostic testing for neurodevelopmental disorders.

## INTRODUCTION

Mendelian disorders causing multiple congenital anomalies and neurodevelopmental dysfunction have an approximately 30-40% chance of getting diagnosed by clinical exome sequencing (ES).^1,2^ Performing ES data reanalysis 1-2 years after the first test can further increase diagnostic yield, but a substantial proportion of patients (50-60%) remain undiagnosed.^3,4^ This is partly due to limitations in the interpretation of identified variants (e.g. variants of uncertain significance) and undiscovered gene-disease relationships. Limitations with ES technology also results in missing certain variant types, such as structural rearrangements, repeat expansions, and non-coding regulatory and deep intronic variants impacting gene expression and splicing.^2^ Characterizing intronic variants that disrupt splicing is especially critical as splice-region mutations cause highly damaging effects on gene products and represent about 10% of all disease causing variants.^5^ Although genome sequencing (GS) can detect non-coding variants, this method alone cannot prove if there is a functional consequence to the nearest transcript. Alternative approaches are therefore necessary to capture pathogenic events causing Mendelian conditions.

RNA sequencing (RNA-seq) is the best approach currently available to detect genome-wide differences in transcript abundance and splicing.^6^ Additionally, this method can identify germline exonic SNVs, indels, and allele specific expression.^7-11^ The utility of RNA-seq in the setting of genomic diagnostic testing has been applied to a selection of neuromuscular^9,10^ and mitochondrial disorders.^11^ The decision to investigate the diagnostic value of RNA-seq in these conditions likely reflects the relative accessibility of affected tissue (i.e. muscle biopsy for neuromuscular disease and fibroblasts from skin biopsy for mitochodriopathy). Complex protocols to derive muscle lineage cells from patient fibroblasts has also been shown to yield testable material that recapitulates the transcriptional diversity of primary muscle tissue.^9^ Effort to circumvent invasive biopsies and complex cell culture protocols for RNA-seq testing was done by testing whole blood of patients with predominately neurological phenotypes and a suspected Mendelian condition.^8^ This approach, however, had limited efficacy with a 7.5% diagnostic yield across all patients.^8^ Therefore, implementing RNA-seq into diagnostic practice is restricted by primary tissues that do not have an ideal transcriptional landscape to test for genes causing phenotypically variable, rare disorders.

Here we demonstrate the utility of RNA-seq on patient derived B-lymphoblastoid cell lines (LCLs) for the diagnosis of patients presenting with multiple congenital anomalies and neurodevelopmental phenotypes. LCLs are made by Epstein-Barr virus (EBV) transformation of mature B lymphocytes and have been broadly used in human genetics research for decades.^12^ LCL transcriptome analysis revealed biologically relevant isoforms of known or predicted candidates of Mendelian genes causing neurodevelopmental phenotypes and/or multiple congenital anomalies (neurodevelopmental Mendelian genes, NMG). Upon establishing a resource of expressed and testable NMGs by LCL RNA-seq, we optimized RNA-seq bioinformatic parameters to enable detection of abnormal splicing events and coding and non-coding pathogenic variants from mRNA. The efficacy of this approach was demonstrated on LCLs from patients with a clinical diagnosis of Cornelia de Lange Syndrome (CdLS [MIM:122470]); a rare autosomal dominant, multisystem disorder caused by pathogenic variants in cohesin complex proteins and characterized by growth and developmental delay, facial dysmorphism, microcephaly, hirsutism, limb anomalies, and congenital heart defects.^13^ Our work establishes LCL RNA-seq as a viable frontline diagnostic tool in neurodevelopmental disorders and as a vital reflexive test when DNA testing is negative.

## METHODS

### Patient samples, LCL preparation, and RNA-seq

All patient samples were obtained with informed consent and with the approval of the internal research ethics board at Children’s Hospital of Philadelphia. Patient samples utilized in this study had a clinical genetics evaluation resulting in a diagnosis of CdLS or related neurodevelopmental disorder. Generation of patient LCLs was performed as previously described.^14^ Briefly, EBV containing supernatant was obtained from EBV producing B95-8 marmoset lymphoblastoid cells cultured in RPMI 1640 medium supplemented with 20% FBS, 2 mM L-glutamine and 1X penicillin-streptomycin (Gibco, Thermo Fisher). Heparinized blood specimens from patients (3-5 ml) was diluted with HBSS (Gibco) and layered onto Ficoll-Plaque Plus (GE) to perform density separation. Collected peripheral blood mononuclear cells were then transduced with EBV containing supernatant. LCL colonies were typically obtained within 2 weeks and subsequently expanded and stored at low passage before downstream analysis. RNA was extracted from cultured LCLs using the miRNeasy Mini Kit (QIAGEN) as per the manufacturer’s instructions. All samples met the required quantity and quality cut-offs with >500 ng of total RNA and an RNA Integrity Number (RIN) ≥9.5. Sequencing was performed at BGI CHOP using non-stranded poly-A selection of mRNA (Illumina TruSeq) followed by paired-end 100 bp sequencing (HiSeq 4000) to a coverage of ≥100 million reads per sample. Our mRNA poly-A library preparation protocol was similar to that of the Genotype-Tissue Expression sequencing project (GTEx)^15^ to ensure consistency between our data and the GTEx control dataset. DNA was extracted using DNeasy kit (QIAGEN) according to manufacturer’s protocol.

### Gene expression analysis for GTEx LCL, blood and brain tissues and CdLS-LCL patient samples

Processed gene and isoform expression data was downloaded from the UCSC Computational Genomics laboratory using STAR alignment and RSEM normalization using hg38 as reference genome and GENCODE v23 gene annotation.^16,17^ In order to harmonize expression data, we processed CdLS-LCL patient samples using the same pipeline: STAR aligner v2.5.4b to align reads to hg38 reference genome and RSEM v1.2.28 to quantify gene expression in terms of FPKM (Fragments Per Kilobase of transcript per Million mapped reads) and isoform expression in terms of TPM (Transcripts Per Million). To determine expression of a genes across all samples, we first filtered our gene list to only protein-coding genes and used a cut-off of mean FPKM >1 and mean coverage >10x to identify expressed protein-coding genes. A curated list of 2541 NMGs (gene list downloaded from publically available GeneDx Autism/ID Xpanded Panel) were annotated with OMIM phenotype identifiers (api.omim.org) and overlapped with the set of expressed protein-coding genes in each dataset to identify expressed NMGs in CdLS (n=1745), GTEx LCLs (n=1706) and GTEx whole blood (n=917). Scripts used to perform principal component analysis, expression scatterplots, bar plots and gene list intersections is available at github.com/komalsrathi/MendelianRNA-seq. Additional plots were generated with GraphPad Prism v5.

### RNA-seq data processing for detecting abnormal splice events

We used paired-end RNA-sequencing data from 15 CdLS-LCL samples to detect abnormal transcript splicing. In addition, raw fastq files containing paired-end RNA-sequencing reads were downloaded from dbGaP accession: phs000424.v7.p2 for GTEx LCL (n=106) and Whole Blood (n=336). Paired-end fastq files from all three datasets were aligned to the hg19 genome using STAR v2.5.4b aligner in two-pass mode. Splice junctions identified from the 1^st^ pass alignment were filtered for mitochondrial junctions and for any unannotated junctions that were supported with less than 5 reads. The filtered splice junctions were concatenated across all samples per dataset and were used as input in order to realign the reads with STAR 2^nd^ pass alignment. Following the 2^nd^ pass alignment, duplicated reads were tagged using Picard MarkDuplicates utility.

The first step in abnormal splice junction discovery was to use the sorted, de-duplicated bam files in order to extract all splice junctions supported by uniquely mapped reads for each sample. Next, read support normalization was done on the resulting splice junctions in order to correct for variability in gene expression and library size. This step was done in order to transform the raw read counts into proportion of reads that support a splice junction compared to all other overlapping junctions. Using the above approach, we found a total of 3534 splice junctions in CdLS samples (n=15) and a total of 13980 splice junctions in GTEx samples (n=442) corresponding to the 15 known CdLS genes.

The next step was to filter the junctions to identify potential deleterious splice events. Abnormal splice events were identified by multiple criteria: 1) those that were seen at a level of at least 5% of canonical junctions, 2) number of reads that support the junction in the entire dataset ≥10, 3) not seen in GTEx normal samples and 4) identified in at most one patient (patient-specific events). The resulting patient-specific abnormal transcript splice events were visualized in the form of sashimi plots that were generated using the R package ggsashimi.

### Variant calling pipeline

Raw RNA-seq reads were aligned to GRCh37/hg19 reference genome using STAR version v2.5.4b.^17^ Picard MarkDuplicate version 2.18.14-0 command was used to remove PCR duplicates. We used GATK HaplotyperCaller v3.6 instead of v4 to achieve higher precision.^9,18^ Variants were called according to GATK best practices for RNA-seq variant calling with minor modifications.^19^ Applying SplitNCigarReads to the bam files reduced overall variant calling sensitivity therefore we excluded this step from the final pipeline. Instead, Filter_reads_with_N_cigar parameter was applied in HaplotypeCaller to remove reads with CIGAR strings (?) containing N operator. Variant calling region was set to exons of the genes (exons of all transcripts) listed in Table S6 and surrounding 10 bp flanking intronic regions. Impact prediction of variants was performed using SnpEff v4.3.^20^ The following filters were applied to the variants called (condition to keep variants listed): 1) quality filter: QD>=2 and FS<30, 2) variant impact filter: Synonymous, missense, or any variants marked as impact “HIGH” by SnpEff, 3) common variants filter: gnomAD maximum subpopulation frequency (AF_PopMax) <0.2%. Coverage statistics were calculated using GATK DepthOfCoverage v3.6 over two bed files: 1) exons of the genes used for variant calling and 2) 10bp flanking intronic regions of those genes. Variants from BAM files were displayed using Alamut (v2.11).

## RESULTS

Using RNA-seq as a diagnostic tool for Mendelian disorders requires extracting mRNA from accessible patient tissue. Determining which tissue to assay is critical as the disease causing gene could be silenced or display tissue specific splicing that obscures data interpretation. To determine if blood is an appropriate specimen type for RNA-seq, we compared its gene expression profile to LCLs using data from the GTEx.^15^ Choosing to compare blood with LCL was done since EBV transformation of B lymphocytes is a relatively easy and reproducible method to generate patient cell lines with very low mutation rates and stable karyotypes.^12,21-23^ We set an expressed gene threshold of >1 FPKM and read depth of >10x across exonic regions. Overall, nearly 2-fold more genes were expressed above this threshold in LCLs (n=10612) compared to blood (n=5617), and of those genes expressed in blood, 93% (5243/5617) were found in LCL, indicating LCLs provide a similar complement of expressed genes and over five-thousand more testable genes (Fig. 1a). We next explored the expression of 2541 curated NMGs (Table S1). Sixty-seven percent of NMGs (1706/2541 genes) were expressed in LCLs above the expressed gene threshold compared to 36% in blood (917/2541 genes) (Table S1). This equaled 1.8x more NMGs that are testable in LCL compared to blood. Additionally, the relative expression of NMGs was higher in LCLs compared to blood (Fig. 1b). These analyses indicate LCLs provide a comprehensive transcriptional landscape to perform RNA-seq based diagnostic testing.

**Fig. 1.**
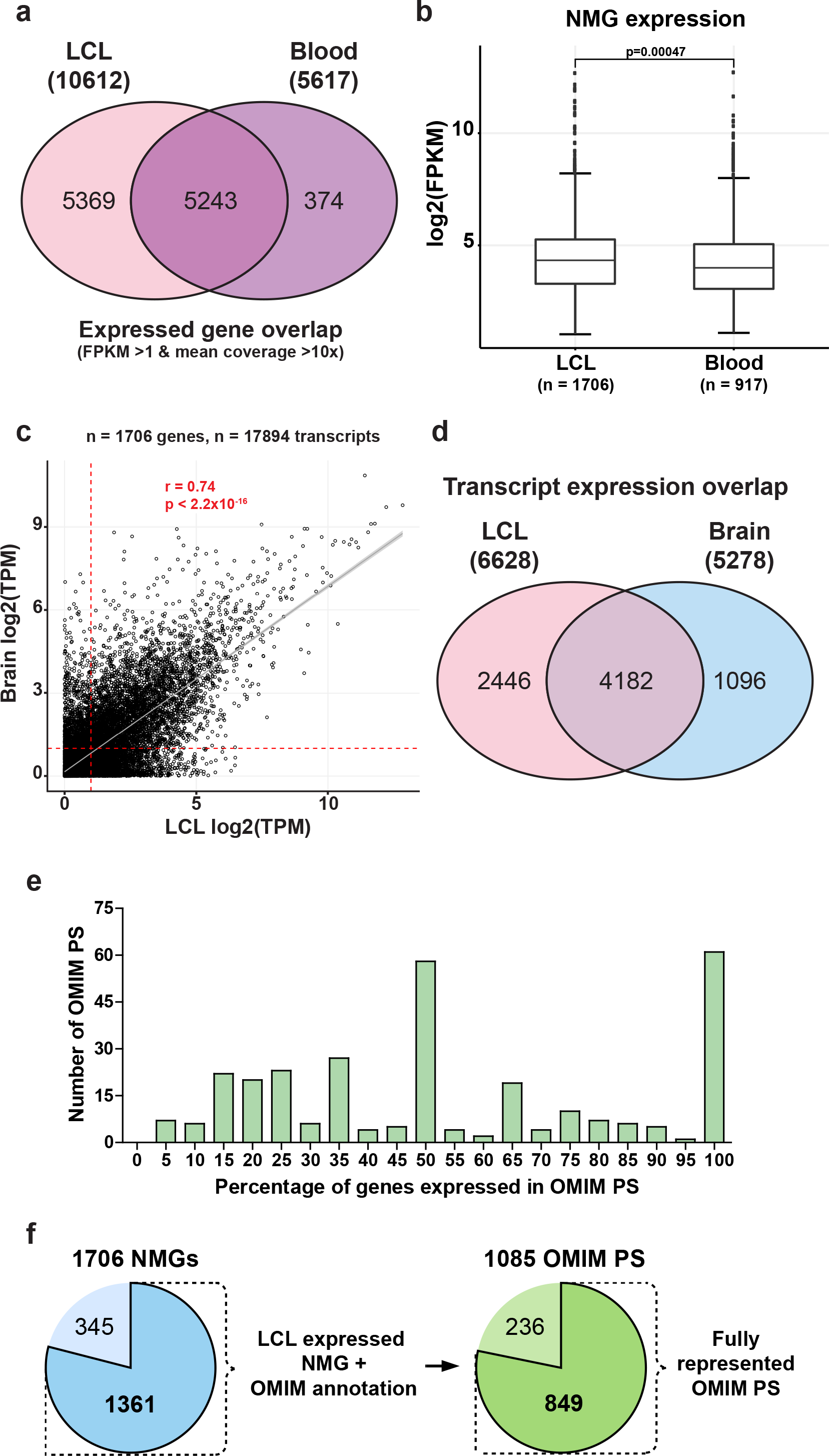
Genes involved in neurodevelopmental Mendelian disorders are expressed in LCLs. **a** Total number and overlap of expressed transcripts in GTEx LCL and blood samples meeting the expressed gene threshold of FPKM >1 and mean coverage >10x. **b** Mean expression (FPKM) of NMGs in LCL (n=1706 genes) and blood (n=917 genes). Data shown as mean ±SEM. **c** Correlation of isoform expression between GTEx LCL and brain. **d** Overlap of transcript isoforms that meet the expressed gene threshold in brain and LCL. **e** Histogram depicting number of OMIM PS with partial or complete gene coverage in LCLs from a total of 297 PS. **f** Charts depicting the number of LCL expressed NMGs (n=1706) that have a documented disease-gene relationship in OMIM (dark blue) and the subsequent breakdown of these genes into OMIM PS (n=1085) with complete (dark green) or partial representation (light green).

Expression of biologically relevant isoforms were determined by comparing to GTEx brain tissue. A total of 17894 transcript isoforms were found in GTEx for the 1706 LCL expressed NMGs (Table S2). Expression of all 17894 transcripts correlated between LCL and brain (r=0.74, p<2.2×10^−16^) (Fig. 1c), but only 37% exceeded the expressed gene threshold (6628/17894), which translates to an observed 3-4 expressed isoforms per gene for LCL. In brain we found similar results with expression of 29% of annotated isoforms (5278/17894) (Fig. 1c). LCL expressed isoforms had 63% overlap with brain (4182/6628) (Fig. 1d), and between these two groups there was 65% overlap for the max expressed transcript (1114/1706). Altogether, there is significant positive correlation in isoform expression between brain and LCLs and overlap in expression of the most biologically relevant transcripts.

Next we investigated what types of genes were represented in the list of 1706 LCL NMGs. GO analysis of molecular function found significant enrichment for genes associated with catalytic activity (p=2.86×10^−65^), nucleotide binding (p=7.3×10^−34^), and chromatin binding (p=3.03×10^−12^) (Table S3). These gene functions were reflected in biological processes that showed enrichment for metabolic processes (p=5.4910-35), mitochondrion organization (1.07×10-22), chromosome organization (p=8.64×10-24), and nervous system development (p=2.18×10-21) (Table S3). Examining GO term enrichment for 835 NMGs that did not meet the expressed gene threshold in LCLs showed enrichment for terms such as ion channel (p=3.72×10^−42^) and neurotransmitter receptor activities (p=1.40×10^−21^) (Table S4). Thus, LCLs are appropriate specimens for diagnostic testing for a large variety of NMGs participating in many core cell processes, but display limitations for testing transcripts with highly restricted neural lineage expression.

A clinical diagnosis for a Mendelian disorder is based on a recognizable pattern of phenotypic features and is typically due to mutation in a single gene. Phenotype-gene relationships are curated into Phenotypic Supersets (PS) in the Online Mendelian Inheritance of Man (OMIM)^24^ (Figure S1). From the 1706 NMGs expressed in LCLs, 1361 were involved in 2008 OMIM PS. After removing duplicate PS, there was a total of 1085 unique PS, of which 788 were represented by one gene and 297 by more than one gene (Table S5). RNA-seq from LCLs detected all genes in 61/297 PS (20.5%) that had more than one disease causing gene (Fig. 1e). Examples of PS with full gene representation included Congenital disorders of glycosylation, type II (PS212066, 17 genes), Mitochondrial complex III deficiency (PS124000, 9 genes), Aicardi-Goutieres syndrome (PS225750, 7 genes), and CdLS (PS122470, 5 genes) (Table S5). The cumulative number of PS with 50-100% of genes represented by RNA-seq was 177/297 (60%) (Fig. 1e). Combining the 788 PS represented by a single gene with 61 PS that have all genes expressed results in complete gene representation in 78% (849/1085) of observed OMIM PS (Fig. 1f). Overall, LCLs express hundreds of single genes that cause neurodevelopmental disorders as well as coexpress many complete sets of genes that comprise OMIM phenotypic series.

We next used this information to develop a diagnostic RNA-seq pipeline for LCL specimens from patients with Mendelian disorders. Our protocol uses transcriptome data and bioinformatic filters for expressed transcripts containing deleterious coding and non-coding variants (+/-10 bp intron) and abnormal splicing events. Pipeline validation was performed on LCLs made from patients with CdLS (CdLS-LCL), since all genes comprising the CdLS PS are expressed in normal LCLs (Fig. 2a; Table S5). The validation cohort consisted of 10 CdLS-LCL specimens with various mutations in four CdLS genes (*NIPBL, SMC1A, SMC3* and *HDAC8*) (Table 1). Gene expression profiles of CdLS-LCLs were compared with GTEx blood, brain and LCL samples. Principle component analysis showed GTEx LCL and CdLS-LCL groups cluster together relative to blood indicating similarity in their global expression profiles (Fig. 2b; Figure S2a). CdLS-LCL expression of 18385 transcript isoforms from 1745 NMGs meeting the expressed gene threshold (1745/2541 NMGs) showed positive correlation with brain (r=0.68, p<2.2×10^−16^) and very strong correlation with GTEx LCL (r=0.96, p<2.2×10^−16^), with 89.9% of genes having the same highest expressed isoform (Fig. 2c; Figure S2b). In line with correlated expression with GTEx LCL, there was 98% similarity in the number of genes captured per OMIM PS (1050/1071) (Figure S2c; Table S5). Thus, the measurable expression landscape for diagnostic testing did not greatly differ between control GTEx LCL and CdLS-LCL groups.

**Table 1.**
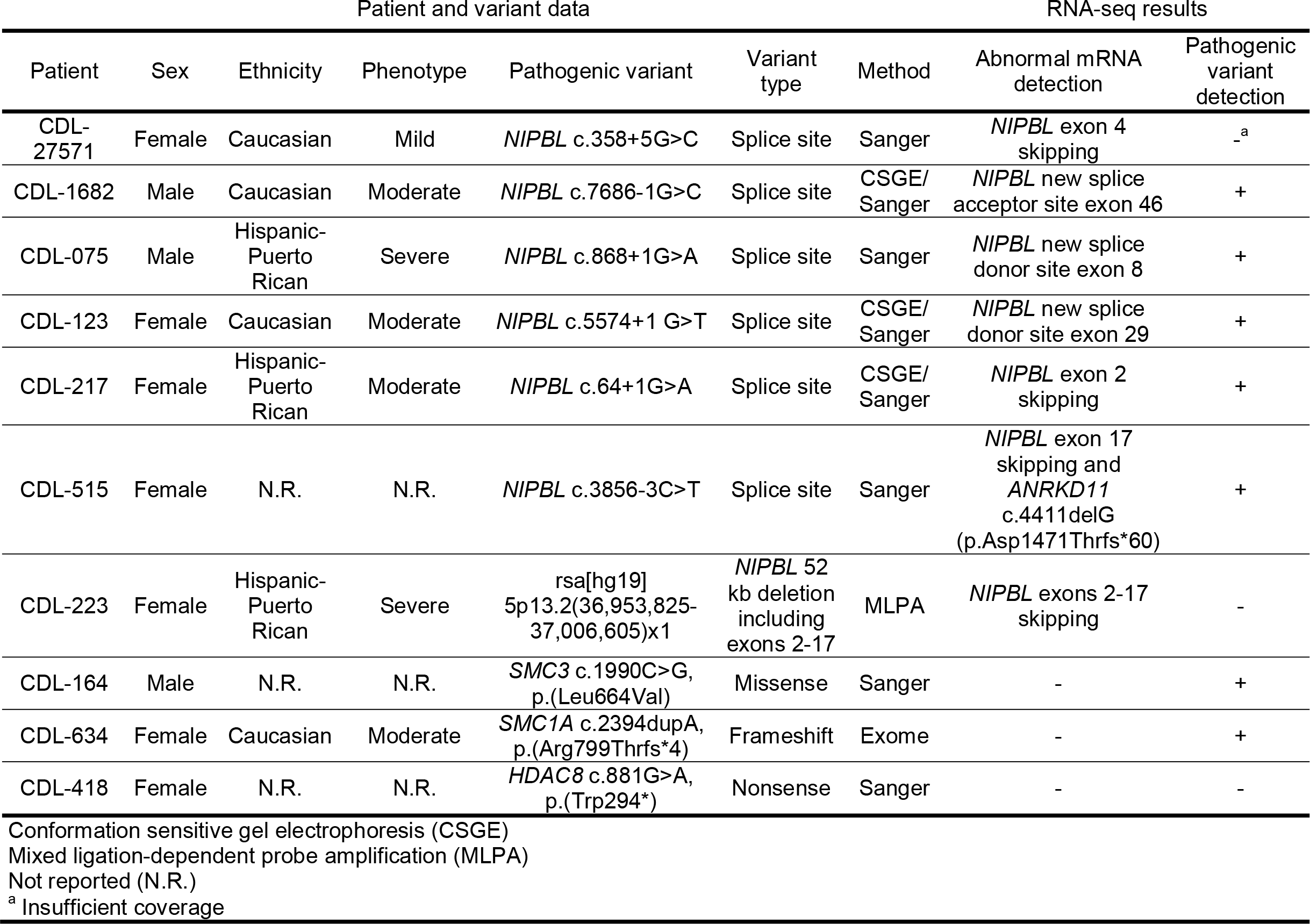
RNA-seq results for CdLS-LCL validation cohort.

| Patient and variant data |  |  |  |  |  |  | RNA-seq results |  |
| --- | --- | --- | --- | --- | --- | --- | --- | --- |
| Patient | Sex | Ethnicity | Phenotype | Pathogenic variant | Variant type | Method | Abnormal mRNA detection | Pathogenic variant detection |
| CDL-27571 | Female | Caucasian | Mild | *NIPBL* c.358+5G>C | Splice site | Sanger | *NIPBL* exon 4 skipping | -<sup>a</sup> |
| CDL-1682 | Male | Caucasian | Moderate | *NIPBL* c.7686-1G>C | Splice site | CSGE/<br>Sanger | *NIPBL* new splice acceptor site exon 46 | + |
| CDL-075 | Male | Hispanic-Puerto Rican | Severe | *NIPBL* c.868+1G>A | Splice site | Sanger | *NIPBL* new splice donor site exon 8 | + |
| CDL-123 | Female | Caucasian | Moderate | *NIPBL* c.5574+1 G>T | Splice site | CSGE/<br>Sanger | *NIPBL* new splice donor site exon 29 | + |
| CDL-217 | Female | Hispanic-Puerto Rican | Moderate | *NIPBL* c.64+1G>A | Splice site | CSGE/<br>Sanger | *NIPBL* exon 2 skipping | + |
| CDL-515 | Female | N.R. | N.R. | *NIPBL* c.3856-3C>T | Splice site | Sanger | *NIPBL* exon 17 skipping and *ANRKD11* c.4411delG (p.Asp1471Thrfs*60) | + |
| CDL-223 | Female | Hispanic-Puerto Rican | Severe | rsa[hg19] 5p13.2(36,953,825-37,006,605)x1 | *NIPBL* 52 kb deletion including exons 2-17 | MLPA | *NIPBL* exons 2-17 skipping | - |
| CDL-164 | Male | N.R. | N.R. | *SMC3* c.1990C>G, p.(Leu664Val) | Missense | Sanger | - | + |
| CDL-634 | Female | Caucasian | Moderate | *SMC1A* c.2394dupA, p.(Arg799Thrfs*4) | Frameshift | Exome | - | + |
| CDL-418 | Female | N.R. | N.R. | *HDAC8* c.881G>A, p.(Trp294*) | Nonsense | Sanger | - | - |

Conformation sensitive gel electrophoresis (CSGE) Mixed ligation-dependent probe amplification (MLPA) Not reported (N.R.) <sup>a</sup> Insufficient coverage

**Fig. 2.**
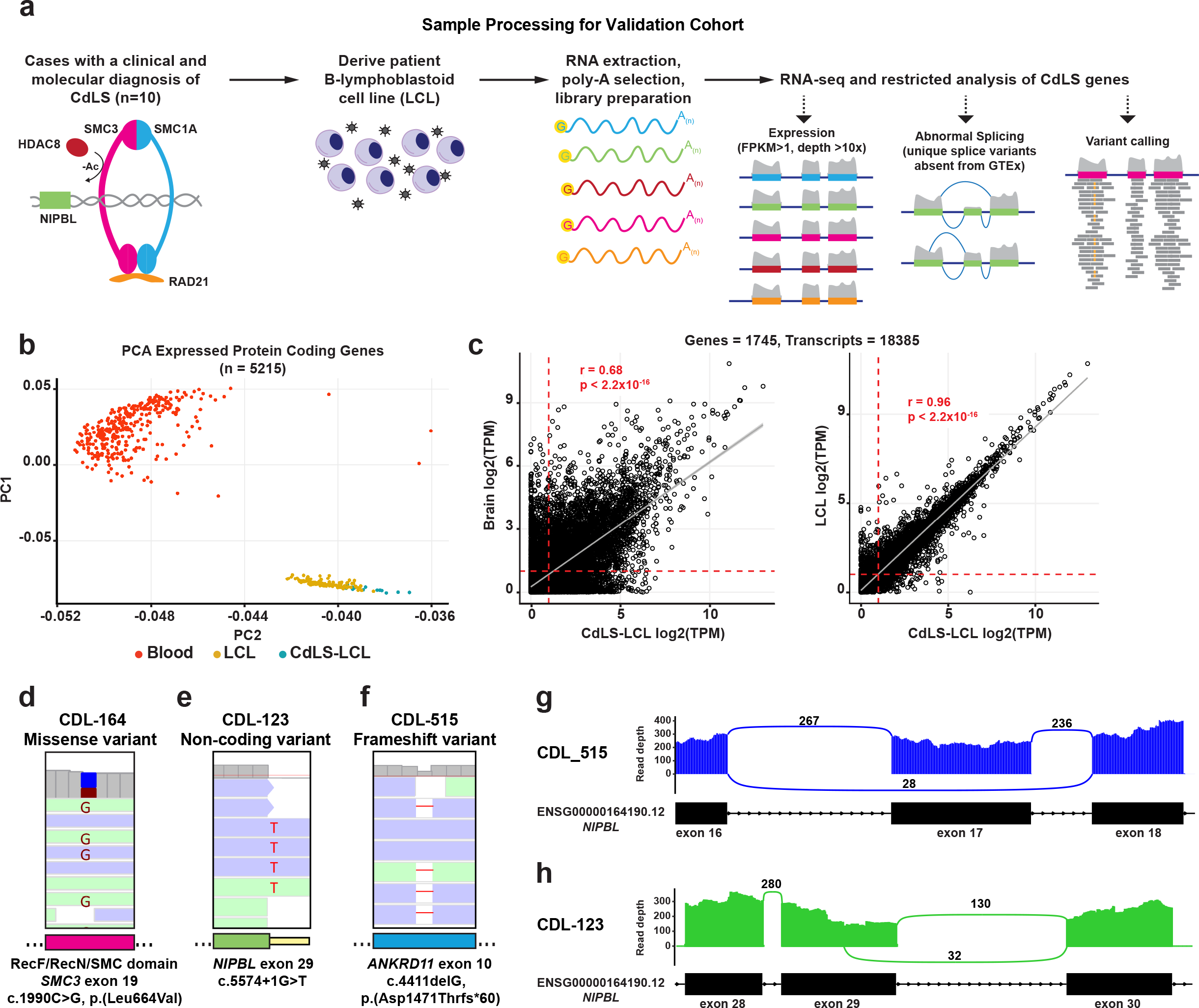
LCL RNA-seq identifies pathogenic variants and abnormal splicing in a validation cohort of CdLS patients. **a** Schematic of sample processing and RNA-seq analysis performed on the validation cohort of patient LCLs. **b** PCA analysis comparing similarity in gene expression profiles between GTEx blood, GTEx LCL to patient LCLs (CdLS-LCL). **c** Correlation between CdLS-LCL and GTEx brain (left panel), and CdLS-LCL and GTEx LCL (right panel) for all annotated transcript isoforms belonging to 1745 NMGs. **d-f** Representative images of RNA-seq reads capturing pathogenic *SMC3* missense variant (**d**), *NIPBL* splice-site variant (**e**), and *ANKRD11* frameshift variant (**f**). **g** Sashimi plots of abnormal splicing events causing premature termination.

Variant calling in CdLS-LCLs was restricted to an RNA-seq panel of 14 genes causing cohesinopathy-related disorders (Table S6; Figure S3). For the highest expressed transcripts, read coverage of >5x was found across 92% of coding base pairs and >2x in 18% of bases at introns (+/-10 bp intron-exon boundary) (Table S7, S8, S9). This allowed detection of pathogenic coding variants in 2/3 samples and pathogenic splice site variants in 5/6 samples (Fig. 2d, 2e, Table 1). An additional 20 unique variant calls required manual review including 16 splice artifacts (e.g. splice region insertion of sequence from the adjacent exon), 2 false positive coding artifacts (e.g. low VAF variant found in multiple samples), and 2 true positive coding variants. Interestingly, one of these true positive variants is a frameshift in *ANKRD11* which causes KBG syndrome^25^ (Fig. 2f). Thus, this patient has a splice region mutation in *NIPBL* (c.3856-3C>T) and an *ANKRD11* p.Asp1471Thrfs*60 mutation expanding their diagnosis to two syndromes with overlapping clinical features.^26^

The other key feature of our pipeline is identifying abnormal splicing events. Fourteen cohesinopathy-related genes were filtered for unique events by comparing within the CdLS-LCL cohort and to GTEx blood and LCL (n = 442) to remove batch effects and normal splicing variation (Figure S4). Abnormal splice products were detected in 7/7 patient LCL with known *NIPBL* splice region mutations or multi-exon deletion (Figure S5; Table 1). Outcomes included exon skipping and utilization of new donor and acceptor splice sites (Fig. 2g, 2h). No false positive splice events were detected with this approach. Overall, our RNA-seq pipeline showed 90% sensitivity for calling pathogenic events (9/10 pathogenic variants and/or abnormal splice products observed).

To test the utility of our approach on unsolved cases, we performed RNA-seq on 5 patients with moderate to severe clinical presentations of a suspected cohesinopathy and non-diagnostic genetic testing (Table 2). Assessment with our 14 gene panel revealed a positive result in 3/5 cases. Patient CDL-679 had severe CdLS presentation and partial skipping of *NIPBL* exons 33 and 34 with premature termination when exon 32 was linked to 34 and a truncated inframe product when spliced to exon 35 (Fig. 3a). Patient CDL-022 had a c.1038G>C, p.Lys346Asn missense variant in exon 6 of *BRD4* (Fig. 3b), a gene recently found to cause CdLS like phenotypes.^27^ Evidence to support variant pathogenicity includes amino acid conservation, deleterious computational predictions (ClinPred and SIFT),^28,29^ absence from the genome aggregation database (gnomAD),^30^ and occurrence in the second bromodomain which has previously been shown to harbor disease causing mutations.^27^ The patient’s mother did not carry this variant and the father was not available to confirm *de novo* status. The third positive patient, CDL-614, was diagnosed with moderate CdLS and had abnormal *NIPBL* splicing with inclusion of a cryptic exon found in intron 21 that is expected to introduce a premature stop codon (Fig. 3c). The *de novo* exon sits within a segmental duplication that has nearly identical homology to two intergenic regions on chromosomes 17 and 18 (Figure S6a, S6b). Interestingly, this patient is one of six children, of which one sibling also has a clinical diagnosis of CdLS and the other four siblings and parents are healthy (Fig. 3d). Locus specific long-range PCR enabled sequencing of the region containing the cryptic exon. This showed a heterozygous c.4560+2069C>T (chr5:37012396C>T) deep-intronic variant in the proband and affected sibling but not the parents (Fig. 3e). This variant was absent in gnomAD and splice site prediction algorithms confirmed it strongly induces formation of a novel 5’splice donor site (Table S10). We suspect parental germline mosaicism for this mutation given there are two affected siblings with apparently the same *de novo* deep intronic variant.

**Table 2.** CdLS-LCL test cohort RNA-seq results for 14 cohesinopathy genes.

| Patient data and prior negative genetic testing |  |  |  |  |  |  |  |  | RNA-seq results |  |  |
| --- | --- | --- | --- | --- | --- | --- | --- | --- | --- | --- | --- |
| Sample | Gender | Ethnicity | Phenotype | Cytogenetics | *NIPBL* | *SMC3* | *SMC1A* | *HDAC8* | Result | TP pass filter variants<sup>a</sup> | FP pass filter variants<sup>a</sup> |
| CDL-679 | Male | N.R. | Severe | Karyotype | Seq | Seq | Seq | - | *NIPBL* exons 33 and 34 skipping | 0 | 3 |
| CDL-614 | Male | Caucasian | Moderate | Chromosome microarray | Seq/MLPA | Seq | Seq | Seq | *NIPBL* cryptic exon inclusion from intron 21 | 0 | 1 |
| CDL-069 | Male | African American | Severe | - | Seq/MLPA | Seq | Seq | Seq | - | 0 | 0 |
| CDL-086 | Male | Caucasian | Severe | - | Seq/MLPA | Seq | Seq | - | - | 0 | 3 |
| CDL-022 | Male | Caucasian | Moderate | - | Seq/MLPA | Seq | Seq | Seq | *BRD4* c.1038G>C, p.(Lys346Asn) | 1 | 1 |
| Gene sequencing (Seq) |  |  |  |  |  |  |  |  |  |  |  |
| Mixed ligation-dependent probe amplification (MLPA) |  |  |  |  |  |  |  |  |  |  |  |
| True positive (TP) |  |  |  |  |  |  |  |  |  |  |  |
| False positive (FP) |  |  |  |  |  |  |  |  |  |  |  |
| <sup>a</sup> Includes coding and splice region variants |  |  |  |  |  |  |  |  |  |  |  |

**Fig. 3.**
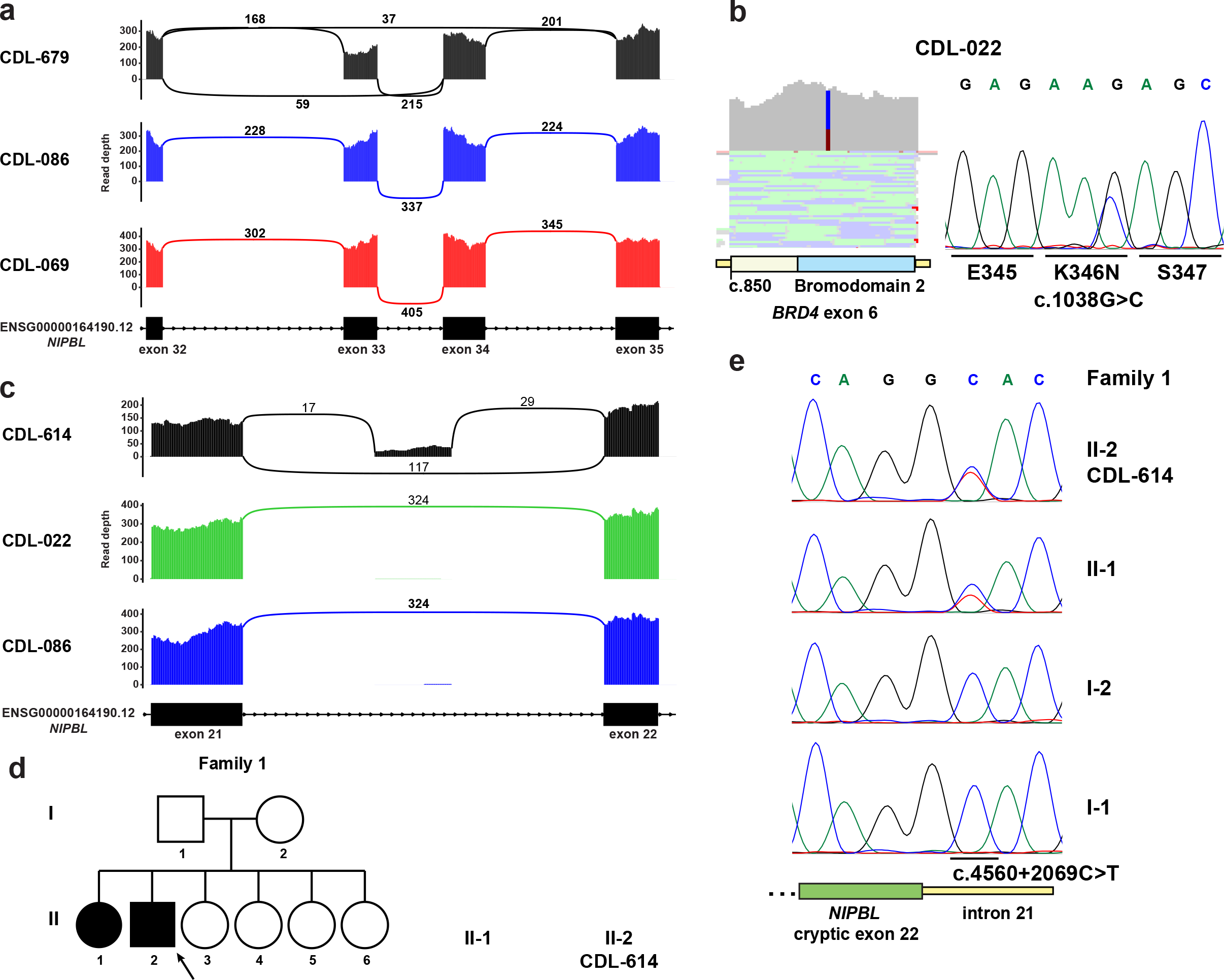
LCL RNA-seq reveals abnormal splicing and pathogenic variants in patients with only clinical diagnoses of cohesinopathy. **a** Sashimi plots from test cohort patients showing abnormal *NIPBL* splicing in patient CDL-679. **b** RNA-seq reads (left) and Sanger confirmation of cDNA (right) of a *BRD4* missense variant. **c** Sashimi plots from test cohort patients showing abnormal *NIPBL* splicing in patient CDL-614. **d** Left panel shows family pedigree for patient CDL-614 (proband indicated by arrow), right panel shows images of proband and affected sibling with CdLS. **e** Locus specific LR-PCR followed by Sanger sequencing of *NIPBL* intron 21 in proband, affected sibling, and parents of family 1.

## DISCUSSION

New approaches are needed to find pathogenic events in Mendelian disorders because conventional testing by gene panel and ES leaves many patients undiagnosed. In this study, we show LCLs made from patient blood share the transcriptional repertoire of brain tissue for a significant proportion of NMGs, enabling testing of over 1000 genetic syndromes. This analysis enabled us to create an important new resource of testable genes by RNA-seq for clinical practice (see Tables S2 and S5). Our analysis shows the LCL transcriptome compared to blood displays elevated NMG expression and has nearly 2-fold more total genes expressed above our set threshold. We suspect the limited transcriptional landscape of terminally differentiated blood cells and the heterogeneity of sampled cell populations^31^ limits the number of testable genes in whole blood. Thus, the large diversity of genes with robust expression make LCLs superior to blood for diagnostic testing. We further found a strong similarity between GTEx LCL control and CdLS-LCL expression datasets. This indicates the act of B cell transformation normalizes gene expression and makes a fairly homogeneous testing background. Therefore, while many genes are noted to be differentially expressed between control LCL and CdLS-LCL,^32^ the milieu of testable transcripts above our expression threshold remained nearly identical. This buffering effect is likely a robust phenomenon given cohesinopathy gene perturbation is known to affect global gene expression profiles.^32-35^ Based on these findings we hypothesize that LCL from other genetic disorders will have a nearly identical testing background to normal LCL, which is important for clinicians and laboratories that need to know which genes can be reliably found by this technique. Despite these observed benefits, the drawback of using LCLs is the time and costs required to establish cell lines, which take about one month to produce sufficient quantities of cells for RNA-seq and long-term storage. Additionally, prolonged culturing can yield genomic artifacts,^36,37^ thus all testing should be done on early passage cell lines which show negligible genetic changes compared to primary tissue.^21,36^

Using a combination of 1) variant calling and 2) abnormal splice pattern detection, we obtained 90% sensitivity for all pathogenic events in the validation cohort. Break down of these two features shows all abnormal splice products were detected without introducing false positives, whereas variant calling missed two SNVs (false negatives), including a non-sense variant in *HDAC8* and a +5 splice region variant in *NIPBL*. Dropout of the *HDAC8* p.Trp294* variant on Xq13.1 is likely due to nonsense mediated decay resulting in allele biased expression. Other studies have utilized allelic imbalance to identify pathogenic events across the transcriptome,^8,9^ however, we find challenges with implementing this strategy in a practical, clinical setting. One difficulty is constraint in variation in haploinsufficient genes,^38-40^ which can result in an absence of heterozygous SNVs in clinically significant genes within individuals. We noticed this phenomenon with *HDAC8* where no coding region variants were detected in our validation cohort. Additionally, we found the *HDAC8* nonsense variant did not significantly alter gene expression (Figure S3). Thus allele imbalances and expression changes can be useful as a screening strategy to prioritize genes carrying pathogenic variants across the transcriptome, but their interpretation should be treated with caution given the absence of these differences does not preclude an actual pathogenic variant is present. The second variant that was not detected was *NIPBL* c.358+5G>C. Our pipeline was designed to retain reads containing exon read-throughs since we suspected disruptive splice region variants to be included in newly mutated mRNA products. This +5 variant was not observed due to lack of coverage at this intronic position, however all pathogenic splice variants from positions ±1-3 were found. Deeper sequencing could be one strategy to boost detection of exon read-through events. Retaining reads carrying intronic variants for analysis of splice altering mutations is especially important if RNA-seq is going to be used before testing by exome or genome sequencing.

We applied our LCL RNA-seq testing strategy on five patients that had gone several years without genetic diagnosis for their clinical presentation of CdLS or related cohesinopathy. This led to identification of two abnormal *NIPBL* splice events and one missense variant in *BRD4*. One of the abnormal *NIPBL* splice events resulted in the inclusion of a cryptic exon and represents a novel mechanism of pathogenic inactivation. The high homology of the region containing the cryptic exon and pathogenic deep intronic variant is expected to have mapping quality issues by genome sequencing. This highlights the strength of RNA-seq for demonstrating the impact of mutations found in regions of high homology. The severe CdLS-like phenotypic presentation of the patient carrying the *BRD4* missense variant in the second bromodomain is in line with previous studies showing *de novo* missense variants in this region causes typical CdLS dysmorphic facial appearance, intellectual disability, and developmental delay.^27^ The similarity in patients with pathogenic variants in *BRD4* and *NIPBL* reflects work showing BRD4 directly interacts with NIPBL to bind enhancers of developmentally important genes.^27^ The two remaining patients that did not receive diagnosis by analyzing 14 CdLS-related genes suggests unknown cohesinopathy genes could be involved. Overall, this work illustrates the large testable transcriptional landscape of patient LCLs for neurodevelopment disorders and shows the specific application of LCL RNA-seq in providing new diagnoses in CdLS patients through detecting defective splicing and pathogenic coding variants.

## Data Availability

All data can be available upon request

## SUPPLEMENTARY INFORMATION

Supplementary Information includes six figures and ten tables and Supplementary Materials and Methods.

## ACKNOWLEDGEMENTS

We would like to thank the families involved in this study and Dr. Nancy Spinner and Deborah McEldrew for their technical support. This work was supported by internal departmental funds.

